# Right ventricular contractility decreases during exercise in patients with non-advanced idiopathic pulmonary fibrosis

**DOI:** 10.1101/2020.05.14.20098327

**Authors:** Sandra de Barros Cobra, Marcelo Palmeira Rodrigues, Felipe Xavier de Melo, Nathali Mireise Costa Ferreira, César Augusto Melo-Silva

## Abstract

**Introduction:** Early right ventricular dysfunction in non-advanced patients with idiopathic pulmonary fibrosis has not been fully elucidated. Thus, we aimed to assess right ventricular functions in idiopathic pulmonary fibrosis patients and controls by speckle-tracking strain echocardiography at rest and peak exercise.

**Methods:** We conducted a cross-sectional study in 20 idiopathic pulmonary fibrosis patients without oxygen use, blood oxygen saturation levels ≥92% at rest, and modified Medical Research Council score ≤3 and enrolled 10 matched controls. Transthoracic echocardiography images were acquired at rest and during a cardiopulmonary exercise test. We analyzed two-dimensional echocardiographic parameters and right ventricular function using the global longitudinal strain assessed by the two-dimensional speckle-tracking technique.

**Results:** In the control group, we found normal values of global longitudinal strain (GLS) at rest and at peak exercise, the latter being much more negative (−23.6±2.2% and -26.8±3.1%, respectively; p<0.001). By contrast, GLS values in the idiopathic pulmonary fibrosis group increased from -21.1±3.8% at rest to -17.0±4.5% at peak exercise (p<0.001). The exercise revealed a difference between the two groups as the mean GLS values moved during peak exercise in opposite directions. Patients with idiopathic pulmonary fibrosis got worse, whereas control patients presented improved right ventricular contractility.

**Conclusions:** Right ventricular dysfunction was unveiled by speckle-tracking echocardiography during exercise in non-advanced idiopathic pulmonary fibrosis patients. We suggest that this reflects an inadequate right ventricular-arterial coupling decreasing the right ventricular longitudinal contraction during exercise in these patients. This parameter may be useful as an early index of suspected pulmonary hypertension.

## Introduction

Idiopathic pulmonary fibrosis (IPF) is a progressive and irreversible chronic lung disease [1]. Pulmonary hypertension (PH) is common in advanced IPF [2,3] and increases the risk of death by approximately threefold [4]. The prevalence varies between 3-86%, but is most commonly 30-50% [5].

Typically, some IPF outcomes are related to PH and the adaptation of the right ventricle (RV) to the afterload changes. In these cases, RV failure components may drive mortality [6]. Despite its critical relevance, RV function remains unclear in these patients, particularly in those without severe hypoxemia at rest. Patients with non-advanced IPF and PH are probably the best study candidates to increase our understanding of “disproportional PH.”

Ventriculoarterial uncoupling is the physiological consequence of failing RV adaptation. The gold standard for assessing ventriculoarterial coupling is invasive, involving cardiac catheterization; however, it is difficult to assess during exercise, as it requires manipulation of the venous return [7]. Two-dimensional transthoracic echocardiography (2D TTE) allows a complementary investigation of RV behaviors in diverse pulmonary disease stages. D’Andrea et al., used RV speckle-tracking echocardiography (STE) to evaluate the global longitudinal strain (GLS), and demonstrated that RV STE is an accurate tool for the evaluation of right ventricular function, is easy to perform, and feasible in various clinical scenarios of RV dysfunction [8].

Evaluation of exercise-induced changes in pulmonary hemodynamics may improve understanding on pulmonary circulation-heart interactions. The analysis of RV function during exercise can predict RV failure in IPF [9]. Minor myocardial dysfunction emerging from abnormal systolic contractility can be accurately and non-invasively measured by STE [10] and may even be detected with minor afterload changes [11].

Data are lacking on RV dysfunction at rest and during exercise in patients at early IPF stages with normal or near-normal oxyhemoglobin peripheral saturation (SpO_2_). D’Andrea et al. described RV dysfunction during exercise using STE [12]; however, they included patients with severe hypoxemia at rest (mean SpO_2_, 83%) and probably patients at advanced IPF stages.

To date, no approved medications are available for treating PH in these patients [13]. Evaluating pulmonary circulation-heart interactions and RV functions through ventricular-vascular coupling is a powerful approach that should improve understanding on the disease. Singh et al. demonstrated that RV-pulmonary artery dissociation could compromise RV contractility in response to increased afterloads during exercise [14]. This increased afterload is probably insufficient to maintain a normal cardiac output and will eventually result in the deterioration of RV–pulmonary artery (PA) coupling at peak exercise.

We hypothesized that increased pulmonary artery pressure and consequent RV dysfunction may occur at early IPF stages as part of its development. We also hypothesized that STE can detect RV function impairments during exercise even in IPF patients without RV dysfunction at rest, and could serve as an early index for suspected PH.

## Methods

This study was approved by the Ethics Committee of School of Medicine - University of Brasilia - Brazil (protocol number CAAE: 71022817.2.0000.5558). All patients signed an informed consent to participate and a consent for publication.

We evaluated 116 consecutive IPF patients between February and December 2017 who were diagnosed according to the criteria of the American Thoracic Society and European Respiratory Society [15]. Treatment for interstitial lung diseases followed in a tertiary care center at the University Hospital of Brasília. Among 116 subjects, we selected 20 for 2D TTE at rest and cardiopulmonary exercise testing (CPET); 2D TTE was performed as per the American Society of Echocardiography guidelines [16].

Inclusion criteria for IPF patients included a Gender-Age-Physiology Index score ≤5, compatible with mild or moderate disease [17], modified Medical Research Council scale [18] score ≤3, and SpO_2_ ≥92% at rest and in room air without oxygen therapy. Patients with locomotor diseases, severe comorbidities (e.g., lung cancer, pulmonary thromboembolism, and stroke), left cardiomyopathy, or combined emphysema and pulmonary fibrosis were excluded.

Controls were recruited from elderly healthy family members and friends of the IPF patients. The controls did not demonstrate respiratory symptoms, had no history of lung diseases, and did not meet the exclusion criteria. We identified 10 controls that met the selection criteria.

The study was approved by the Ethics Committee of School of Medicine - University of Brasilia - Brazil (protocol number CAAE: 71022817.2.0000.5558), and written informed consent was obtained from all subjects. The procedures in this research were in accordance with the ethical standards of the Helsinki Declaration of 1975, as revised in 1983.

## Transthoracic doppler echocardiography at rest

The baseline resting echocardiogram before CPET was performed using a Vivid I (GE Healthcare, Milwaukee, WI, USA). Final values were obtained after averaging over three cardiac cycles. Conventional echocardiographic parameters were acquired to evaluate RV functions. The modified Bernoulli equation was applied to calculate systolic pulmonary artery pressure (sPAP) from tricuspid regurgitation. The formula mPAP = 0.61 × sPAP + 2 mmHg was used [19], assuming that sPAP equaled the right systolic ventricular pressure in the absence of RV pulmonary stenosis or outflow tract obstruction [20]. The estimated right atrial pressure was based on the inferior vena cava collapsibility index, that was added to the sPAP [16, 21]. Images were adjusted for better RV free wall delimitation and STE analysis.

## RV function assessment by 2D TTE and STE

The STE technique [22] was applied to images using the EchoPac software (v. 201; General Electric, Vingmed, Horten, Norway) offline. The beginning and end of the RV systole were defined using the event-timing feature of this software to evaluate RV GLS. Regions of interest were marked manually, and RV free wall edges were adjusted to enable adequate tracing of basal, medial, and apical portions of the RV myocardium. This was performed at both rest and peak exercise; images were adjusted for better delimitation of the RV free wall endocardium (Figs 1 and 2). The average of three segments defined the measurement results. The systolic peak longitudinal strain reflecting muscle fiber contraction was expressed as a percentage and has a negative value [22]. Current reference values for global RV free wall STE suggest that values ≤-20% are normal [16].

**Fig 1.**
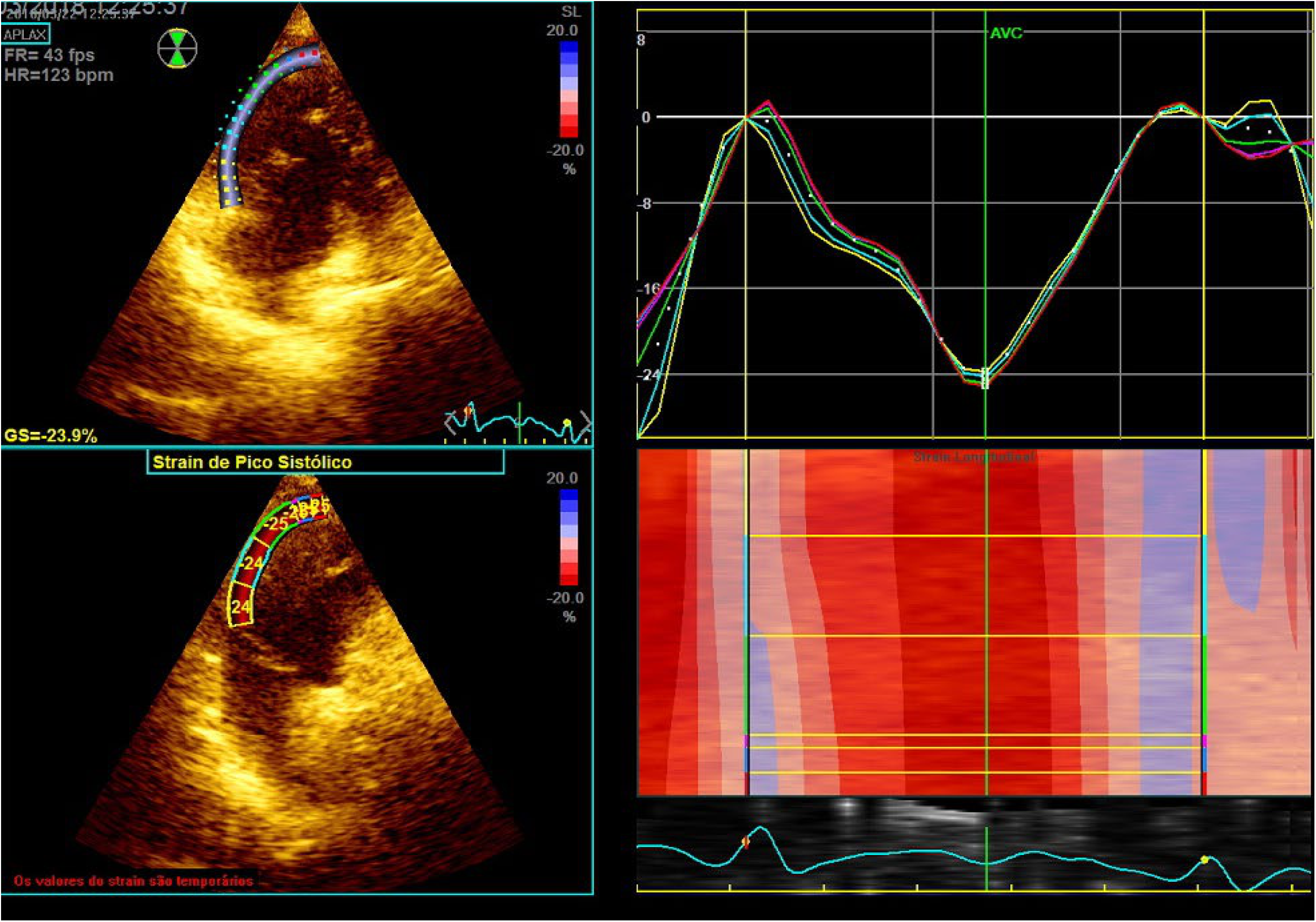
Normal global longitudinal strain (GLS%) obtained at rest in a patient with IPF.

**Fig 2.**
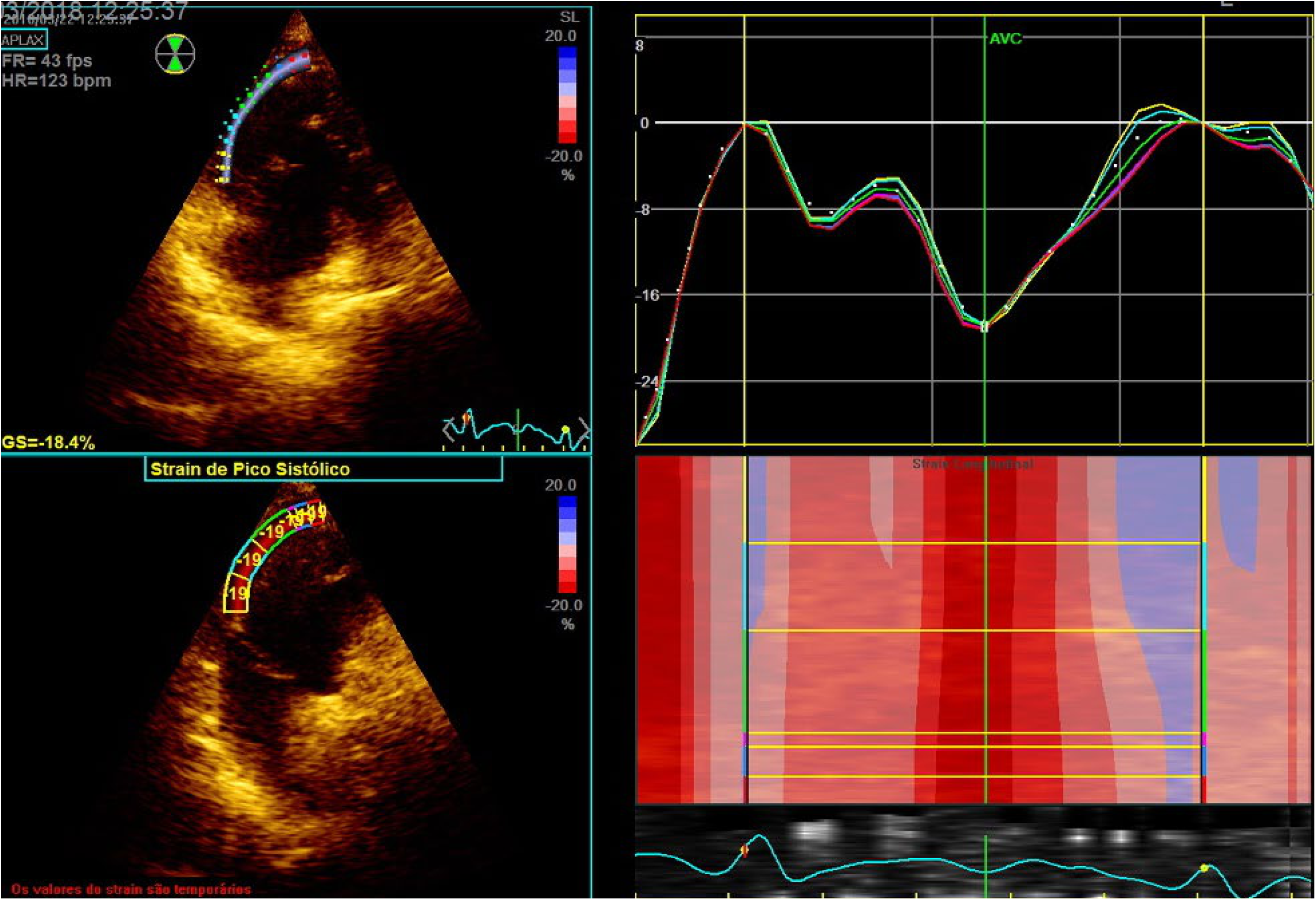
Abnormal global longitudinal strain (GLS%) obtained at peak exercise in the same patient with IPF.

## CPET with simultaneous echocardiography

All participants were tested using an incremental ramp protocol. A cycle ergometer (CG-04; Inbramed^®^, Porto Alegre, RS, Brazil) was used with a progressively increasing load. The protocol consisted of pedaling for 1 min at 60 rotations/min without load (0 W). The load was then increased in 5-W steps from 5 to 30 W/min. As per the American Thoracic Society recommendations [23], we recorded the dyspnea level, exercise capacity, maximum predicted value of oxygen extraction (V’O_2_), and patient age. These parameters were analyzed during an incremental load period of 8-12 min.

Cardiovascular and ventilatory variables were assessed once all expiratory gas measurements at each breath had been documented (Quark PFT; Cosmed, Rome, Italy). Pulse oximetry findings (Ipod^®^; Nonin Medical, Inc., Plymouth, MN, USA), heart rates, and electrocardiographic parameters were continuously recorded. V’O_2_ released CO_2_ (V’CO_2_), CO_2_ pressures, volume expired per minute (V’E), and final expiratory O_2_ were registered every 15 sec. The auscultatory method was employed to record systolic and diastolic blood pressures at all load increases. The gas exchange method allowed non-invasive estimation of the anaerobic threshold [23]. Equal scales, which include the V-slope technique (slope of the V’O_2_ versus V’CO_2_ graph), were used to analyze this parameter. To determine the performance of ventilatory equivalents (V’E/V’CO_2_ and V’E/V’O_2_) and their final expiratory pressures [23], the ventilatory method was used, validating the V-slope technique.

The Borg effort perception scale [24] was applied to evaluate muscle fatigue sensations and dyspnea during CPET, and echocardiography images were obtained to identify tricuspid regurgitation better [16]. Simultaneous images were stored every minute for the offline sPAP analysis.

During the recovery period, tricuspid regurgitation spectral traces were recorded for 3-4 minutes.

## Statistical analysis

Normal distribution was confirmed by the Kolmogorov-Smirnov test for all continuous variables. The data are presented as the mean±standard deviation, and 95% confidence intervals (95% CIs) are shown. Student’s *t*-test for independent samples was used to compare means of continuous variables. Categorical variables are presented as percentages. We used the chi-squared test to compare proportions. Analyses between GLS measures and CPET variables was calculated using Pearson correlations. Data were analyzed using SPSS for Mac OS X^©^ (v.25.0.0; SPSS, Inc., Chicago, IL, USA). Results were considered statistically significant at p<0.05.

## Data

The datasets used and/or analyzed during the current study are available from the corresponding author on reasonable request.

## Results

The study population comprised 30 participants: 10 controls and 20 patients with IPF. Table 1 summarizes the demographic, echocardiographic, and pulmonary function test data at baseline. The two groups did not differ in terms of sex (p=0.605) and age (p=0.219).

**Table 1.** Demographic and echocardiographic data of the IPF and control groups at rest.

| Variable <sup>a</sup> | IPF<sup>b</sup><br>(n=20) | Control<br>(n=10) | 95% CI of<br>difference | P |
| --- | --- | --- | --- | --- |
| Age (year) | 72.3 ± 8.6 | 68.5 ± 5.2 | -9.8 to 2.3 | 0.219 |
| Sex (Female/Male) | 10/10 | 6/4 |  | 0.619 |
| SpO<sub>2</sub> rest (%) | 93.2 ± 1.6 | 94.1 ± 1.4 | -0.03 to 2.43 | 0.056 |
| FVC% | 67.1 ± 15.2 | - |  | - |
| FEV<sub>1</sub>% | 74.3 ± 11.7 | - |  | - |
| D<sub>L</sub>CO% | 53.9 ± 19.0 | - | - | - |
| LA (mL/m<sup>2</sup>) | 20.8 ± 6.2 | 14.7 ± 4.4 | -10.5 to -1.4 | 0.011 |
| RA (mL/m<sup>2</sup>) | 15.5 ± 5.8 | 12.5 ± 5.6 | -7.5 to 1.6 | 0.199 |
| RV FAC (%) | 46.8 ± 9.9 | 62.0 ± 7.6 | 7.8 to 22.4 | <0.001 |
| RV TAPSE (mm) | 20 ± 3 | 22 ± 2 | 0.4 to 5.1 | 0.012 |
| TDI RV Sm peak (cm/s) | 12 ± 2 | 13 ± 2 | 1.0 to 1.6 | 0.695 |
| sPAP (mmHg) | 33.4 ± 10.8 | 22.8 ± 7.1 | -18.4 to -2.8 | 0.009 |
| mPAP (mmHg) | 21.8 ± 6.2 | 15.4 ± 4.5 | -10.9 to -1.7 | 0.008 |
| E/E' average | 7.7 ± 1.9 | 7.2 ± 1.3 | -1.9 to 0.8 | 0.458 |

<sup>a</sup>D<sub>L</sub>CO% carbon monoxide diffusing capacity of the lung, E/E' mitral E/E' ratio, FEV<sub>1</sub>% forced expiratory volume in 1 s as a percentage of the predicted value, FVC% forced vital capacity as a percentage of the predicted value, IPF idiopathic pulmonary fibrosis, LA left atrium, mPAP mean pulmonary artery pressure, RA right atrium, RV FAC right ventricle fractional area change, sPAP systolic pulmonary artery pressure, SpO<sub>2</sub> peripheral oxyhemoglobin saturation, TAPSE tricuspid annular plane systolic excursion, TDI RV Sm peak tissue Doppler imaging of lateral tricuspid systolic annulus velocity
<sup>b</sup>Data are presented as the mean ± standard deviation

As seen in Table 1, regarding conventional TTE parameters, the following variables presented significant differences between groups: RV fractional area (p<0.001), left atrial volume (p=0.011), and tricuspid annular plane systolic excursion (p=0.012). However, the mean values and all the individuals’ values in those three variables were within normal range. There were no significant differences between the study groups in terms of right atrial volume (p=0.199), Mitral E/E’ (mitral valve E velocity by mitral annular E’ velocity ratio) values for left ventricular end diastolic pressure (p=0.458) and peak tissue Doppler imaging values of the lateral tricuspid systolic annulus velocity (p=0.695).

As seen in Table 2, the resting mPAP difference, determined by echocardiography, was higher in the IPF group than in controls (p=0.008). As expected, mPAP values increased during exercise in both groups. However, peak exercise mPAP values were even higher in IPF patients (p=0.015).

**Table 2.** Echocardiographic data at rest and their corresponding exercise parameters.

| Variable<sup>a</sup> | IPF<sup>b</sup> | Control<sup>b</sup> | 95% CI of |  |
| --- | --- | --- | --- | --- |
|  | (n=20) | (n=10) | difference | p |
| mPAP (at rest, mmHg) | 21.8 ± 6.2 | 15.4 ± 4.5 | -10.9 to -1.7 | 0.008 |
| mPAP (peak exercise, mmHg) | 42.4 ± 16.0 | 28.0 ± 9.8 | -25.7 to -2.9 | 0.015 |
| mPAP (peak – rest, mmHg) | 20.6 ± 11.8 | 12.6 ± 7.6 | -16.4 to 0.4 | 0.001 |
| GLS% (at rest) | -21.1 ± 3.8 | -23.6 ± 2.1 | -5.1 to 0.2 | 0.068 |
| GLS% (peak) | -17.0 ± 4.5 | -26.8 ± 3.1 | -13.1 to -6.5 | <0.001 |
| GLS% (peak) - GLS% (at rest) | 4.1 | -3.2 | -10.2 to -4.4 | <0.001 |

<sup>a</sup>GLS global longitudinal strain, *IPF* idiopathic pulmonary fibrosis, *mPAP* mean pulmonary artery pressure
<sup>b</sup>Data are presented as the mean ± standard deviation

Echocardiographic data revealed that in the control group, all participants had normal GLS values at rest and at peak exercise, the latter demonstrating considerably greater negativity (−23.6±2.16% and −26.8±3.1%, respectively, 95% CI: 2.0 to 4.4, p<0.001). This indicates that the RV contractility had increased overall. Differently, in the entire IPF group, GLS values changed from −21.1±3.8% at rest to −17.0±4.5% at peak exercise (95% CI: 2.1 to 6.1, p<0.001), which indicates that the RV contractility mean had decreased. During exercise, only three IPF patients presented slightly improved RV contractility; from rest to peak exercise, their values changed as follows: −14.5 to −19.6, −16.0 to −18.0, and −16.8 to −18.0.

IPF patients could be divided into two groups: (1) normal GLS at rest with decreased RV contractility during exercise; (2) abnormal GLS at rest that also worsened with exercise. In this first subgroup, the 12 IPF patients (60%) with normal GLS values at rest (−23.49±2.91%) declined during exercise (−18.23±4.62%, 95% CI: 3.5 to 6.9, p<0.001). In this second subgroup, eight IPF patients (40%) had abnormal GLS values at rest (−17.70±1.85%) that worsened at peak exercise (−15.27±4.14%, 95% CI: −2.3 to 7.1, p=0.266), however, this difference did not achieve statistical significance.

Comparing IPF patients with controls, GLS values were only significantly different at peak exercise (controls: −17.0±4.5%, IPF: −26.8±3.1%, 95% CI: −13.1 to −6.5, p<0.001; Table 2). The difference in GLS at rest between IPF patients and controls was not statistically significant (−21.1±3.8% and −23.6±2.1%, respectively, 95% CI: −5.1 to 0.2, p<0.068). Importantly, the mean GLS values of the two groups moved in opposite directions during exercise (Fig 3). On STE analyses, RV contractility worsened in IPF patients, and improved in controls. GLS differences between the two groups were also more apparent at peak exercise.

**Fig 3.**
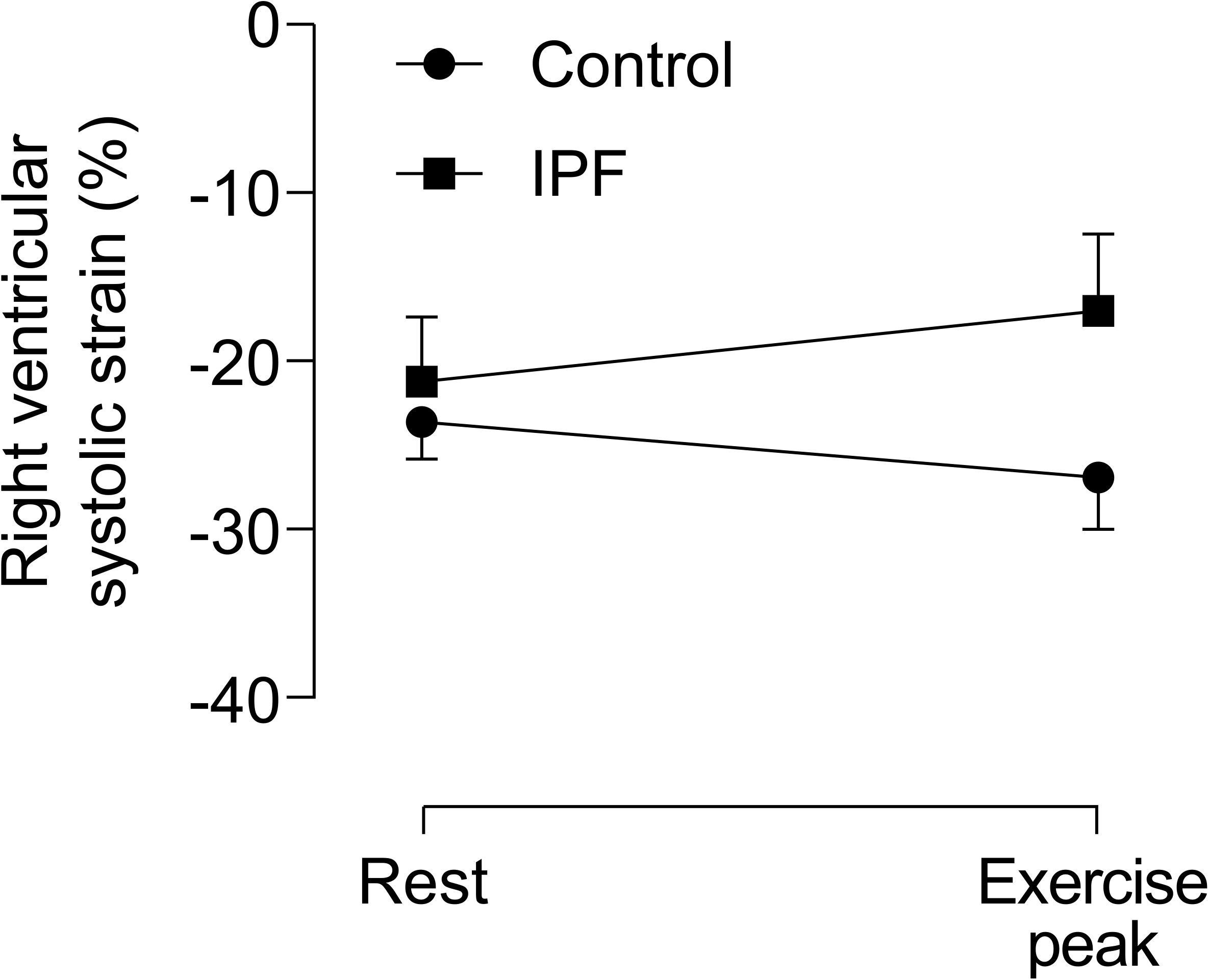
Right ventricular systolic strain shifts from rest to peak exercise in control participants and patients with IPF.

We also performed a correlation analysis between the main CPET variables and GLS values at rest and at peak exercise. Most Pearson correlation coefficients demonstrated a weak-to-moderate correlation (Table 3); mainly those that referred to GLS at peak exercise were statistically significant.

**Table 3.**
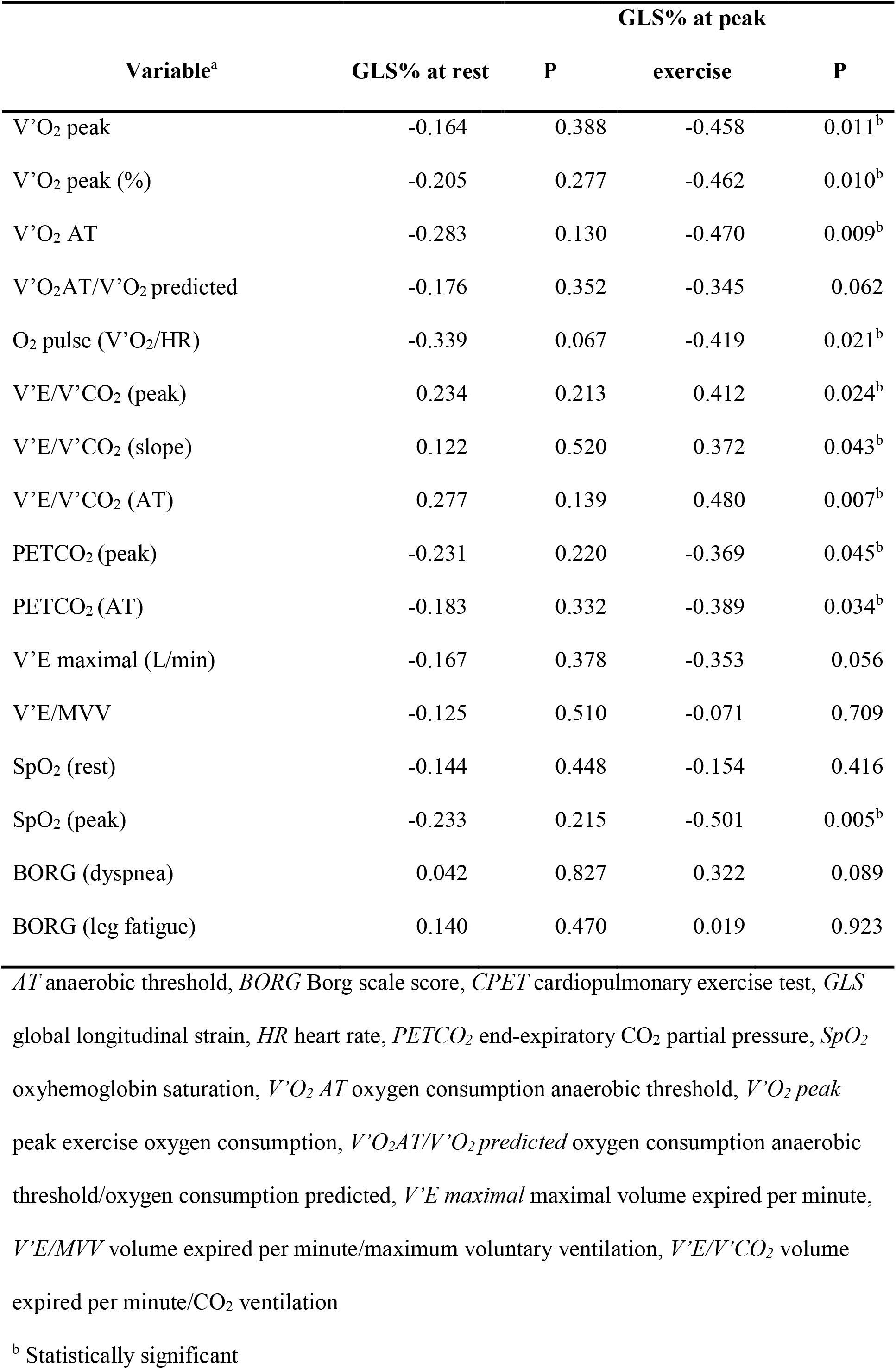
Pearson correlation coefficients between CPET data and GLS values.

| Variable<sup>a</sup> | GLS% at rest | P | GLS% at peak |  |
| --- | --- | --- | --- | --- |
|  |  |  | exercise | P |
| V'O<sub>2</sub> peak | -0.164 | 0.388 | -0.458 | 0.011<sup>b</sup> |
| V'O<sub>2</sub> peak (%) | -0.205 | 0.277 | -0.462 | 0.010<sup>b</sup> |
| V'O<sub>2</sub> AT | -0.283 | 0.130 | -0.470 | 0.009<sup>b</sup> |
| V'O<sub>2</sub>AT/V'O<sub>2</sub> predicted | -0.176 | 0.352 | -0.345 | 0.062 |
| O<sub>2</sub> pulse (V'O<sub>2</sub>/HR) | -0.339 | 0.067 | -0.419 | 0.021<sup>b</sup> |
| V'E/V'CO<sub>2</sub> (peak) | 0.234 | 0.213 | 0.412 | 0.024<sup>b</sup> |
| V'E/V'CO<sub>2</sub> (slope) | 0.122 | 0.520 | 0.372 | 0.043<sup>b</sup> |
| V'E/V'CO<sub>2</sub> (AT) | 0.277 | 0.139 | 0.480 | 0.007<sup>b</sup> |
| PETCO<sub>2</sub> (peak) | -0.231 | 0.220 | -0.369 | 0.045<sup>b</sup> |
| PETCO<sub>2</sub> (AT) | -0.183 | 0.332 | -0.389 | 0.034<sup>b</sup> |
| V'E maximal (L/min) | -0.167 | 0.378 | -0.353 | 0.056 |
| V'E/MVV | -0.125 | 0.510 | -0.071 | 0.709 |
| SpO<sub>2</sub> (rest) | -0.144 | 0.448 | -0.154 | 0.416 |
| SpO<sub>2</sub> (peak) | -0.233 | 0.215 | -0.501 | 0.005<sup>b</sup> |
| BORG (dyspnea) | 0.042 | 0.827 | 0.322 | 0.089 |
| BORG (leg fatigue) | 0.140 | 0.470 | 0.019 | 0.923 |

*AT* anaerobic threshold, *BORG* Borg scale score, *CPET* cardiopulmonary exercise test, *GLS* global longitudinal strain, *HR* heart rate, *PETCO<sub>2</sub>* end-expiratory CO<sub>2</sub> partial pressure, *SpO<sub>2</sub>* oxyhemoglobin saturation, *V'O<sub>2</sub> AT* oxygen consumption anaerobic threshold, *V'O<sub>2</sub> peak* peak exercise oxygen consumption, *V'O<sub>2</sub>AT/V'O<sub>2</sub> predicted* oxygen consumption anaerobic threshold/oxygen consumption predicted, *V'E maximal* maximal volume expired per minute,
$V'E/MVV$ volume expired per minute/maximum voluntary ventilation, $V'E/V'CO_2$ volume expired per minute/ $CO_2$ ventilation
<sup>b</sup> Statistically significant

## Discussion

CPET in combination with STE was suitable for studying RV function in IPF patients. Our findings demonstrated that GLS measured by STE during exercise was capable of unveiling systolic RV dysfunction in non-advanced IPF patients, although RV function measurements at rest by conventional echocardiography were similar between IPF and control groups. Exercise-induced changes in STE may provide an early index for suspected PH in non-advanced IPF stages.

In this study, we found that as measured by STE, the RV function in IPF patients failed to increase appropriately during exercise. In contrast, the RV contractility in controls improved to more negative GLS values. Exercise results in pulmonary hemodynamic changes, elevating the pulmonary artery pressure due to an increase in cardiac output [25]. In healthy subjects, passive distension of the pulmonary circulation and vasodilation-mediated flow changes accommodate this cardiac output with modest mPAP increases and decreased pulmonary vascular resistance [26]. Thus, the RV adapts to increased afterload by increasing contractility.

Our main finding is that IPF patients at non-advanced stages already have an impaired RV contractile reserve. The failure to improve RV contractility in response to a progressive rise in afterload probably results from poor RV–PA coupling at peak exercise. We speculate that GLS STE values at rest do not predict the success or failure to improve RV contractility during exertion, i.e., to uncover contractility changes, it must be measured during exercise.

Unmasking pulmonary vascular disease by raising the cardiac output to demonstrate an increase in pulmonary resistance is a logical idea. In contrast to left ventricular fibers, the predominant longitudinal orientation of RV fibers may minimize the mechanisms that preserve RV systolic function during increased afterload [7]. Furthermore, a thinner RV wall is capable of accommodating an increased preload; however, it is inadequate for greatly increased PA pressures [27]. This may explain the RV dysfunction at peak exercise detected by GLS STE in non-advanced IPF patients.

Additionally, we observed a correlation lack between CPET parameters and STE measurements at rest in the IPF group. However, we found a moderate correlation between STE RV functions and most parameters at peak exercise, supporting the importance of STE measurements during exercise for understanding patients’ physiological changes. A notable and significant correlation was detected between GLS and SpO_2_ during exercise. This indicates subclinical hypoxia as a possible cause for PH and RV dysfunction, even in non-advanced, non-hypoxemic patients at rest, and may be due to SpO_2_ decreases during exercise and/or sleep [28].

Several studies attempted to uncover pulmonary vascular disease by redistributing the flow or increasing the cardiac output utilizing echocardiography with exercise stress [29] or dobutamine administration [30]. Nowadays, these approaches have poor applicability and low reliability. The present study suggests that non-advanced IPF patients have associated RV dysfunction with increased mPAP during exercise. As PH progresses, RV synchrony is lost. Intraventricular asynchrony may be present even in early disease stages [31]. Thus, RV function can be temporarily suppressed at peak exercise, returning to baseline values at rest.

We also compared RV function and pulmonary hemodynamic responses in control subjects and found that exercise stress was associated with a lower mPAP slope, and improved RV contractility. Only a few studies assessed changes in RV function during exercise in healthy subjects. Exercise echocardiography was compared with exercise cardiac magnetic resonance imaging and invasive pressure measurements. They suggested that echocardiography during exercise represents an efficient screening tool to identify RV function changes and pulmonary vascular disease [9].

The current study has some limitations. First, 2D TTE was used to non-invasively assess intracardiac pressure instead of direct measurements using catheterization; however, hemodynamic invasive measurements are not appropriate for patients without advanced IPF. Second, at present there is no specific software for RV STE analysis, and, for this study, we used the software that measures LV function.

There are also questions regarding the interpretation of our findings. First, GLS STE during exercise may offer an early warning marker of global RV dysfunction and may predict the outcome or functional capacity in IPF patients better than STE at rest. Second, as abnormalities in RV functions and PH are present at early disease stages, and are not exclusively caused by hypoxia, other factors may contribute to the development of PH and RV dysfunction.

## Conclusion

Exercise echocardiography may assess RV function in non-advanced IPF patients. Exercise unmasks RV dysfunction, which decreases when an acute overload likely promotes RV-PA uncoupling. Patients with impaired RV function should receive greater attention as this parameter may be an early index of suspected pulmonary hypertension and may indicate poorer prognosis.

## Data Availability

Yes - all data are fully available without restriction

## Declarations

### Authors’ contributions

SBC analyzed and interpreted patient data and was a major contributor to the manuscript preparation. MPR reviewed all data and made substantial contributions to the conception and design of the work. FXM acquired and analyzed CPET data. NMCF selected all patients. CAMS acquired and analyzed the data and revised the work. All authors have read and approved the final manuscript.

## Acknowledgments

The authors especially want to thank Lauro Casqueiro Vianna, PhD, from NeuroVASQ-Integrative Physiology Laboratory (University of Brasília) for providing the ultrasound equipment to execute the exams. We would like to thank Editage for English language editing.

